# International Randomized Controlled Trial Evaluating Changes in Oral Health in Smokers after Switching to Combustion-Free Nicotine Delivery Systems: SMILE Study protocol

**DOI:** 10.1101/2020.12.12.20240143

**Authors:** Gianluca Conte, Sebastiano Antonio Pacino, Salvatore Urso, Pasquale Caponnetto, Francesca Benfatto, Doris Greiling, Eugenio Pedullà, Luigi Generali, Ugo Consolo, Vittorio Checchi, Jan Kowalski, Maciej Nowak, Renata Górska, Amaliya Amaliya, Iain Chapple, Mike Milward, Robert Maclure, Riccardo Polosa, on behalf of the SMILE collaborators

**Affiliations:** Department of General Surgery and Medical-Surgical Specialties, University of Catania, Catania, Italy; Centre for the Prevention and Treatment of Tobacco Addiction (CPCT) Teaching Hospital “Policlinico - V. Emanuele”, University of Catania, Catania, Italy; Addendo srl, Dental Clinic, Catania, Italy; Center of Excellence for the Acceleration of HArm Reduction (CoEHAR), University of Catania, Catania, Italy; Department of Clinical and Experimental Medicine, University of Catania, Catania, Italy; Department of Surgery, Medicine, Dentistry and Morphological Sciences with Transplant Surgery, Oncology and Regenerative Medicine Relevance (CHIMOMO), University of Modena and Reggio Emilia, Modena, Italy; Department of Periodontology, Medical University of Warsaw, Warsaw, Poland; R Maclure Clinical Research, C/: Pedro Salinas No 7, Los Torres 54, Arboleas, 04660, SPAIN; Forschungsdock GmbH, Eidelstedter Weg 1, 22869 Schenefeld, Germany; Universitas Padjadjaran - Dental Faculty, Jalan Sekeloa Selatan no 1, Bandung 40132, West Java – Indonesia; Periodontal Research Group, The University of Birmingham & Birmingham Community Healthcare Trust, 5 Mill Pool Way, Edgbaston, Birmingham B5 7EG; Department of Educational Sciences, University of Catania, Catania, Italy

**Keywords:** Electronic cigarettes, Heated tobacco products, Tobacco Harm Reduction, Smoking, Gingivitis, MGI, Oral Health, Tooth stain, Periodontitis, dental plaque imaging, dental discoloration

## Abstract

Although the well-known detrimental effects of conventional cigarette smoking on oral health, there are still lack of evidences about the impact of less harmful alternatives (such as electronic cigarettes or heat not burn products), especially in young smokers with clinical absence of signs of periodontitis.

SMILE will be a prospective, multicenter, interventional, open label, randomized, controlled, three parallel-arms study assessing oral health parameters and teeth appearance of 18 months duration.

This study aims to compare short- and long-term impact on oral health between smokers continuing with conventional cigarette smoking, those switching to combustion-free nicotine delivery systems (C-F NDS), and never-smokers by objectively evaluating changes in gingival response, as a proxy for periodontal/gingival.

The total number of participants in the study planned of the trial is 606 (505 regular smokers and 101 never-smokers).

Regular smokers not intending to quit will be randomized in the ratio 1:4 either in continuing to smoke commercially manufactured conventional cigarettes (n = 101; Study Arm A) or switching to C-F NDS (n = 404; Study Arm B), never-smokers will be assigned in Arm C (n= 101).

The primary outcome will be to assess and compare the percentage mean change in Modified Gingival Index (MGI) score between Baseline and 18 months follow-up between the Study Arms A and B.

Secondary outcomes include the assessment of within- and between-group (Arm A, Arm B and Arm C) variations from baseline to 18 months follow-up of several endpoints, such as MGI, Tooth Stains Assessment, Dental Discolorations, Plaque Score Imaging, Oral Health Quality of Life (OHQOL) assessment and EuroQoL Visual Analog Scale (EQ VAS – QoL) assessment.

Patient recruitment will start in January 2021 and enrolment is expected to be completed by June 2021.

This will be the first study determining overall oral health impact of using CF-NDS in smokers without sign of periodontitis. Data from this study will provide valuable insights into the overall potential of C-F NDS to reduce the risk of periodontal diseases.

## BACKGROUND

Periodontal diseases, commonly known as gingivitis and periodontitis, arise in response to accumulations of oral bacteria in the gingival sulcus and are characterized by tissue changes in the periodontium occurring as part of the inflammatory process.^(1)^ Gingivitis is characterized by inflammation such as redness, swelling or bleeding on gentle provocation of the gingival sulcus, whereas periodontitis exhibits increased probing depth, clinical attachment loss and radiographic alveolar bone loss, reflecting the destructive aspects of the disease process.^(2,3)^

Periodontal disease is a well-known independent risk factor for cardiovascular disease ^(4,5)^ and reducing gingivitis and periodontitis is likely to have an overall positive impact on overall human health.

There are multiple risk factors for periodontal diseases, but smoking is considered to be an important independent risk factor related to the development and severity of the disease.^(6-9)^ Depending on the definition of disease and the exposure to smoking, the risk to develop destructive periodontal disease is 5-to 20-fold elevated for smokers compared to non-smokers ^(10)^. Active (as well as passive) smokers appear to be prone to more severe periodontal manifestations - even after adjusting for age, education level, history of diabetes, BMI, alcohol consumption, perceived mental stress and oral hygiene levels ^(11)^.

Besides periodontal diseases, cigarette smoking can cause visible dental manifestations including tooth discoloration and tobacco stains, the intensity of which mainly depends on smoking duration and frequency ^(12)^. In a large cross-sectional study in the Uk ^(13)^, smokers were more likely to report tooth discoloration and being dissatisfied with their own tooth colour compared to non-smokers. Dissatisfaction with teeth appearance (due to tooth discoloration and tobacco stains) is often perceived as a significant social problem for smokers. ^(14-15)^

Although it is clear that abstaining from smoking will have beneficial effects on oral health and teeth appearance, most smokers are reluctant to seek formal treatment for stopping smoking with the vast majority making attempts to quit without assistance ^(16,17)^. Consequently, the need for novel and more efficient approaches is required.

Combustion-free technologies for nicotine delivery such as e-cigarettes (ECs) and heated tobacco products (HTPs) are substituting conventional cigarettes globally ^(18)^ and are thought to be less harmful alternative to tobacco smoking ^(19-21)^. However, there are no long-term studies assessing the impact of these new technologies on oral health and teeth appearance.

## METHODS

SMILE is an international, open label, randomized, controlled study of 18 months duration designed to assess whether cigarette smokers switching to combustion free-nicotine delivery systems (C-F NDS) will undergo measurable improvements in oral health parameters and teeth appearance as a consequence of avoiding exposure to cigarette smoke. Specifically, it is intended to assess differences in study endpoints of oral health parameters and dental aesthetics at all study assessment times between participants switching to C-F NDS, participants who continue to smoke conventional cigarettes and participants who have never smoked.

### Study Population

The inclusion and exclusion criteria are summarized in Table 1. Participants will be never-smokers and regular smokers of conventional cigarettes with a clinical absence of signs of periodontitis. They will be a minimum of 18 years of age and any gender included.

**TABLE 1:**
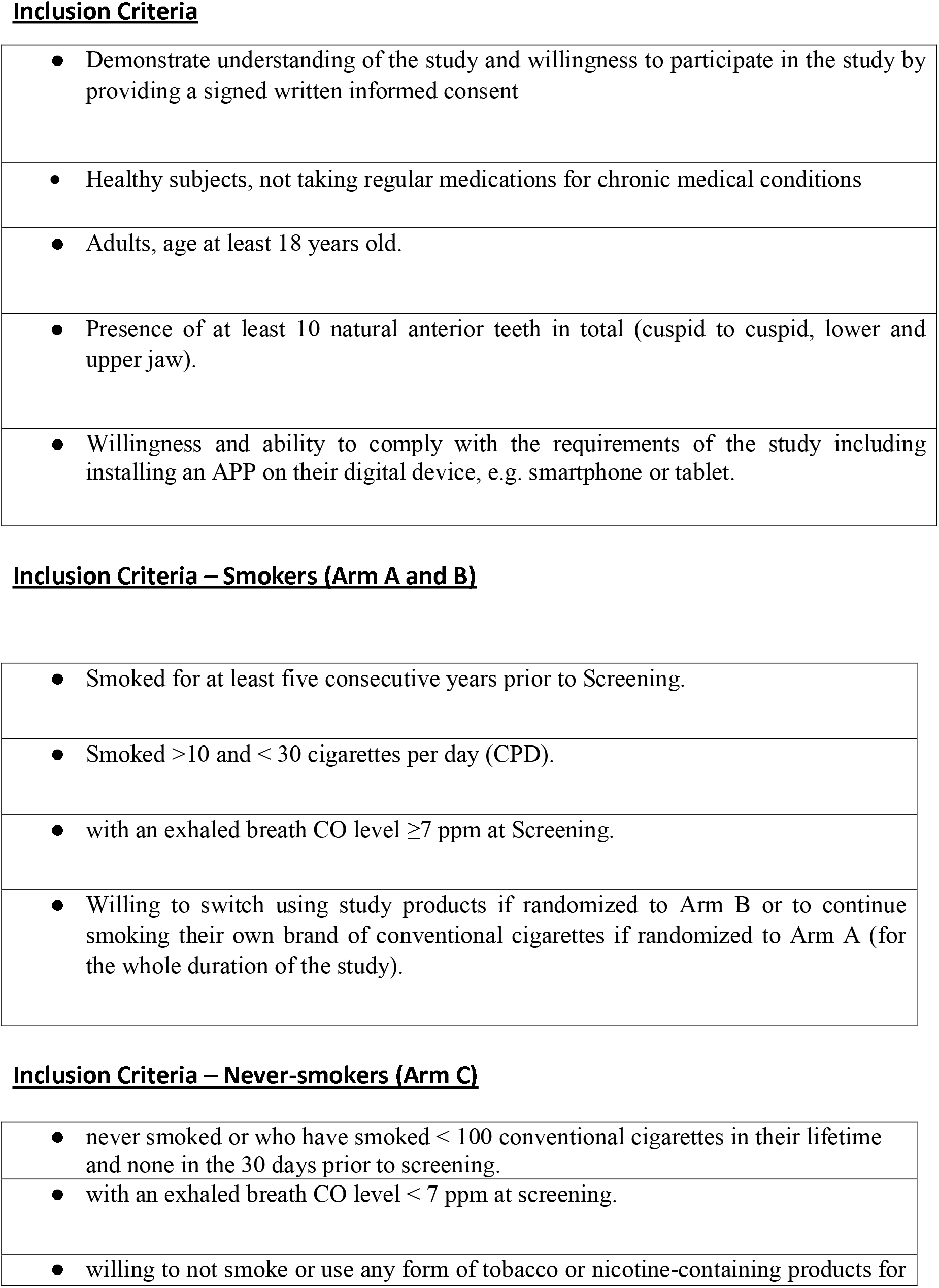
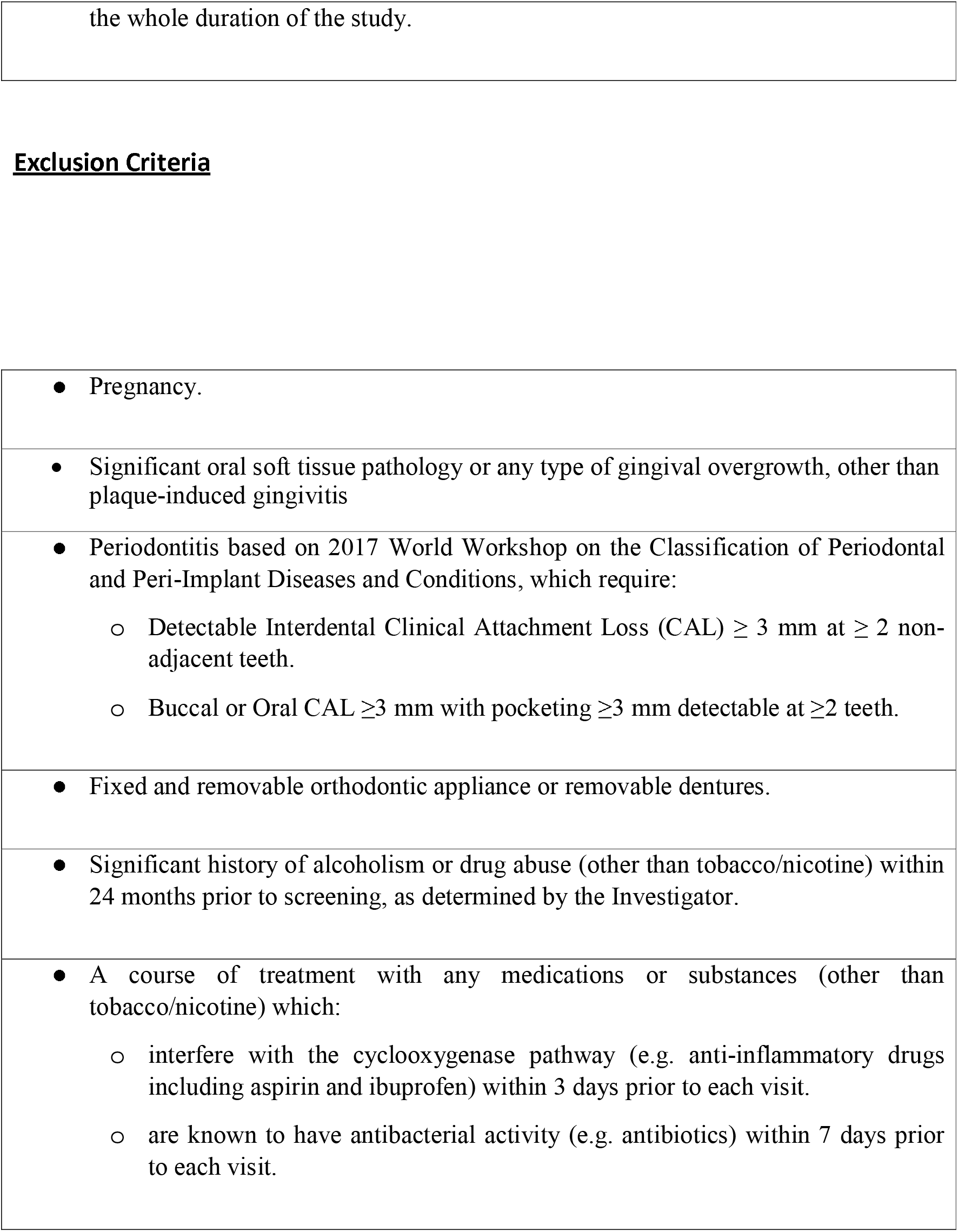
PARTICIPANTS ELIGIBILITY CRITERIA

**Inclusion Criteria**
|  |
| --- |
| • Demonstrate understanding of the study and willingness to participate in the study by providing a signed written informed consent |
| • Healthy subjects, not taking regular medications for chronic medical conditions |
| • Adults, age at least 18 years old. |
| • Presence of at least 10 natural anterior teeth in total (cuspid to cuspid, lower and upper jaw). |
| • Willingness and ability to comply with the requirements of the study including installing an APP on their digital device, e.g. smartphone or tablet. |
| **Inclusion Criteria – Smokers (Arm A and B)** |
| • Smoked for at least five consecutive years prior to Screening. |
| • Smoked >10 and < 30 cigarettes per day (CPD). |
| • with an exhaled breath CO level $\geq 7$ ppm at Screening. |
| • Willing to switch using study products if randomized to Arm B or to continue smoking their own brand of conventional cigarettes if randomized to Arm A (for the whole duration of the study). |
| **Inclusion Criteria – Never-smokers (Arm C)** |
| • never smoked or who have smoked < 100 conventional cigarettes in their lifetime and none in the 30 days prior to screening. |
| • with an exhaled breath CO level < 7 ppm at screening. |
| • willing to not smoke or use any form of tobacco or nicotine-containing products for |
| **Exclusion Criteria** |
| ● Pregnancy. |
| ● Significant oral soft tissue pathology or any type of gingival overgrowth, other than plaque-induced gingivitis |
| ● Periodontitis based on 2017 World Workshop on the Classification of Periodontal and Peri-Implant Diseases and Conditions, which require:<br>○ Detectable Interdental Clinical Attachment Loss (CAL) $\geq 3$ mm at $\geq 2$ non-adjacent teeth.<br>○ Buccal or Oral CAL $\geq 3$ mm with pocketing $\geq 3$ mm detectable at $\geq 2$ teeth. |
| ● Fixed and removable orthodontic appliance or removable dentures. |
| ● Significant history of alcoholism or drug abuse (other than tobacco/nicotine) within 24 months prior to screening, as determined by the Investigator. |
| ● A course of treatment with any medications or substances (other than tobacco/nicotine) which:<br>○ interfere with the cyclooxygenase pathway (e.g. anti-inflammatory drugs including aspirin and ibuprofen) within 3 days prior to each visit.<br>○ are known to have antibacterial activity (e.g. antibiotics) within 7 days prior to each visit. |

505 regular cigarette smokers (>10/day for at least 5 years) will be considered for inclusion. Smoking status will be verified by an exhaled CO measurement (exhaled CO ≥7 ppm).

101 never-smokers (or who have smoked <100 conventional cigarettes in their lifetime and none in the 30 days prior to screening) will be considered for the inclusion. Never smoking status will be verified using exhaled CO measurement (exhaled CO < 7 ppm).

### Ethical Approval

The study will be conducted according to the Principles of Good Clinical Practice (GCP) and Declaration of Helsinki. All six local Ethics Committees reviewed and approved the study and - where appropriate - translated relevant documentation (informed consent form, patient’s information sheet, etc). The trial is registered with ClinicalTrails.gov Identifier: NCT04649645

### Study Design

The study design flow of SMILE is illustrated in Figure 1. All participants will attend a total of seven clinics visits: Day -28 to Day -1 - Screening; Day 0 - Enrollment and Randomization (Visit 0); Day 14 (+/-3 days) - Baseline Visit (Visit 1); Day 90 (+/-5 days) - Visit 2; Day 180 (+/-7 days) - Visit 3; Day 360 (+/-10 days) - Visit 4; Day 540 (+/-10 days) - Visit 5.

**FIGURE 1:**
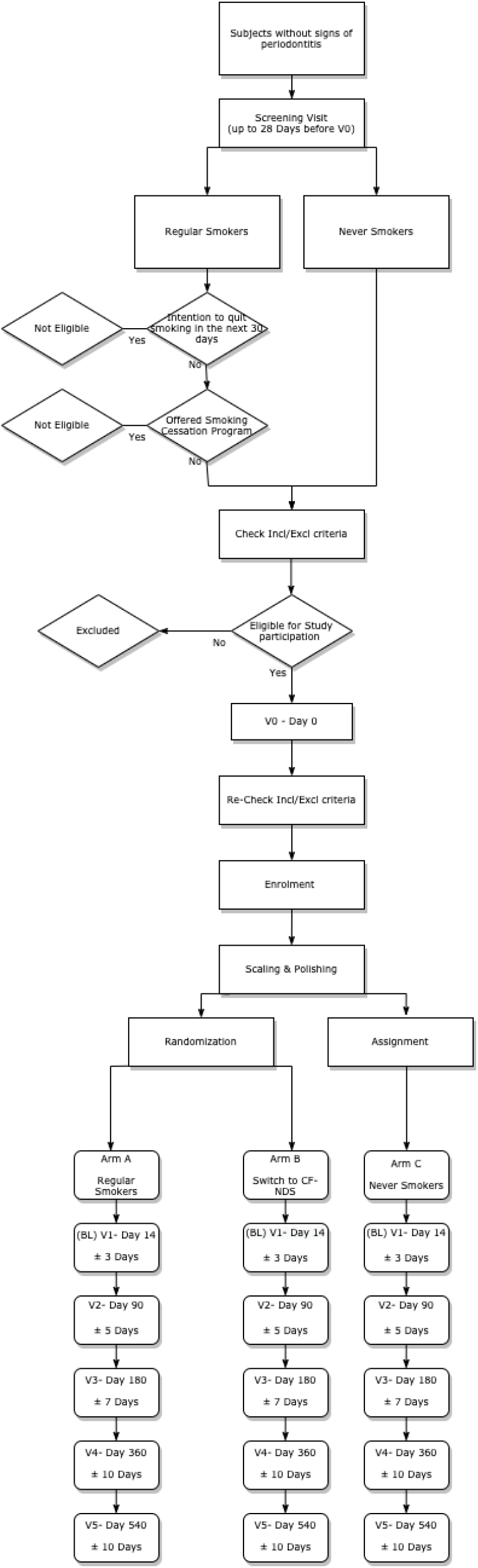
STUDY DESIGN FLOW OF SMILE

Screening visit will be performed within 28 days prior to Visit 0. Procedures conducted at the Screening Visit including socio-demographic data, medical and oral history, smoking history and a screening for periodontal disease, defined according the guidelines of the 2017 World Workshop on the Classification of Periodontal and Peri-Implant Diseases and Conditions ^(22)^. Smokers will be offered smoking cessation program as per local guidelines.

Within 28 days of the screening visit, eligible participants will be invited to attend a second study visit (Visit 0). The inclusion and exclusion criteria will be verified again. Before randomization to one arm, all smokers will be offered a second free smoking cessation program according to standard local guidelines and depending on the local availability of anti-smoking services. Those who refuse to participate in the program will be eligible for recruitment into the study. Smokers will be reminded of the risks associated with smoking and nicotine consumption prior to enrolment onto the study and that they are free to voluntarily quit smoking/nicotine and/or withdraw from the study at any time. Participants will be undergoing a series of assessment (see Table 2a, Table 2b, Table 2c) and a dental scaling and polishing procedures to remove calculus, plaque and stain. Regular cigarette smokers will be randomized 1:4 ratio either continuing to smoke commercially manufactured conventional cigarettes (n = 101; Study Arm A) or switching to C-F NDS (n = 404; Study Arm B). Never Smokers will be assigned to Arm C (n= 101).

**Table 2a:** Study Schedule of Procedures (Arm A – Regular smokers)

| **Procedure** | **Screening**<br><up to 28 days before V0> | **Enrolment**<br>**(V0)**<br>Day 1 | **Baseline**<br>**(V1)**<br>Day 14 $\pm 3$ | **Week 12**<br>**(V2)**<br>Day 90 $\pm 5$ | **Week 24**<br>**(V3)**<br>Day 180 $\pm 7$ | **Week 52**<br>**(V4)**<br>Day 360 $\pm 10$ | **Week 104**<br>**(V5)**<br>Day 540 $\pm 10$ |
| --- | --- | --- | --- | --- | --- | --- | --- |
| Informed Consent | **X** |  |  |  |  |  |  |
| Socio-demographic data | **X** |  |  |  |  |  |  |
| Smoking Hx | **X** |  |  |  |  |  |  |
| Intention to Quit Smoking | **X** | **X** |  |  |  |  |  |
| Smoking Cessation Advice | **X** | **X** | **X** | **X** | **X** | **X** | **X** |
| Cigarette Consumption Check | **X** | **X** | **X** | **X** | **X** | **X** | **X** |
| Exhaled CO | **X** | **X** | **X** | **X** | **X** | **X** | **X** |
| Interest to switch to C-F NDS | **X** |  |  |  |  |  |  |
| FTND | **X** |  |  |  |  |  |  |
| Medical Hx | **X** |  |  |  |  |  |  |
| Assessment of Pregnancy Status | **X** | **X** |  | **X** | **X** | **X** | **X** |
| Oral hygiene Hx | **X** | **X** | **X** | **X** | **X** | **X** | **X** |
| Periodontal Examination | **X** |  |  |  |  |  |  |
| Eligibility Criteria Check | **X** | **X** |  |  |  |  |  |
| Randomization |  | **X** |  |  |  |  |  |
| Tracker APP installation |  | **X** |  |  |  |  |  |
| OHQOL questionnaire |  | **X** | **X** | **X** | **X** | **X** | **X** |
| EQ VAS – QoL questionnaire |  | **X** | **X** | **X** | **X** | **X** | **X** |
| Vital Signs (BP, HR) |  | **X** | **X** | **X** | **X** | **X** | **X** |
| Weight, Height, BMI |  | **X** |  | **X** | **X** | **X** | **X** |
| MGI |  | **X** | **X** | **X** | **X** | **X** | **X** |
| Plaque Score Imaging |  | **X** | **X** | **X** | **X** | **X** | **X** |
| Tooth Stain Assessment |  | **X** | **X** | **X** | **X** | **X** | **X** |
| Dental Discoloration |  | **X** | **X** | **X** | **X** | **X** | **X** |
| Scaling & Polishing |  | **X** |  |  |  |  | **X** |
| Safety reporting (AE and SAE)/e-CRF |  | **X** | **X** | **X** | **X** | **X** | **X** |

**Table 2b:** Study Schedule of Procedures (Arm B – switching to CF-NDS)

| Procedure | Screening<br><up to 28 days before V0> | Enrolment<br>(V0)<br>Day 1 | Baseline<br>(V1)<br>Day 14 ±3 | Week 12<br>(V2)<br>Day 90 ±5 | Week 24<br>(V3)<br>Day 180 ±7 | Week 52<br>(V4)<br>Day 360 ±10 | Week 104<br>(V5)<br>Day 540 ±10 |
| --- | --- | --- | --- | --- | --- | --- | --- |
| Informed Consent | X |  |  |  |  |  |  |
| Socio-demographic data | X |  |  |  |  |  |  |
| Smoking Hx | X |  |  |  |  |  |  |
| Intention to Quit Smoking | X | X |  |  |  |  |  |
| Smoking Cessation Advice | X | X | X | X | X | X | X |
| Cigarette Consumption Check | X | X | X | X | X | X | X |
| Exhaled CO | X | X | X | X | X | X | X |
| Interest to switch to C-F NDS | X |  |  |  |  |  |  |
| FTND | X |  |  |  |  |  |  |
| Medical Hx | X |  |  |  |  |  |  |
| Assessment of Pregnancy Status | X | X |  | X | X | X | X |
| Oral hygiene Hx | X | X | X | X | X | X | X |
| Periodontal Examination | X |  |  |  |  |  |  |
| Eligibility Criteria Check | X | X |  |  |  |  |  |
| Randomization |  | X |  |  |  |  |  |
| Familiarization with C-F NDS |  | X |  |  |  |  |  |
| Tracker APP installation |  | X |  |  |  |  |  |
| OHQOL questionnaire |  | X | X | X | X | X | X |
| EQ VAS – QoL questionnaire |  | X | X | X | X | X | X |

| Procedure | Screening | Enrolment<br>(V0)<br>Day 1 | Baseline<br>(V1)<br>Day 14 ±3 | Week<br>12<br>(V2)<br>Day 90<br>± 5 | Week<br>24<br>(V3)<br>Day<br>180 ±7 | Week<br>52<br>(V4)<br>Day<br>360 ±<br>10 | Week<br>104<br>(V5)<br>Day<br>540<br>±10 |
| --- | --- | --- | --- | --- | --- | --- | --- |
| Vital Signs (BP, HR) |  | X | X | X | X | X | X |
| Weight, Height, BMI |  | X |  | X | X | X | X |
| MGI |  | X | X | X | X | X | X |
| Plaque Score Imaging |  | X | X | X | X | X | X |
| Tooth Stain Assessment |  | X | X | X | X | X | X |
| Dental Discoloration |  | X | X | X | X | X | X |
| Scaling & Polishing |  | X |  |  |  |  | X |
| Switch to C-F NDS |  |  | X |  |  |  |  |
| Safety reporting (AE and SAE)/e-CRF |  | X | X | X | X | X | X |
| Product Count Use |  |  |  | X | X | X | X |

| Procedure | Screen | Enrolment<br>(V0) | Wk<br>2<br>(V1) | Wk<br>4 | Wk<br>8 | Wk<br>12<br>(V2) | Wk<br>20 | Wk<br>24<br>(V3) | Wk<br>32 | Wk<br>40 | Wk<br>48 | Wk<br>52<br>(V4) | Wk<br>60 | Wk<br>68 | Wk<br>76 | Wk<br>84 | Wk<br>92 | Wk<br>100 | Wk<br>104<br>(V5) |
| --- | --- | --- | --- | --- | --- | --- | --- | --- | --- | --- | --- | --- | --- | --- | --- | --- | --- | --- | --- |
| Dispense C-F NDS Device (+ 2 wks supply of either tobacco sticks or e-liquid) |  |  | X |  |  |  |  |  |  |  |  |  |  |  |  |  |  |  |  |
| Dispense 4 wks supply of either tobacco sticks or e-liquid |  |  |  | X | X |  | X |  |  |  | X |  |  |  |  |  |  | X |  |
| Dispense 2 x 4 wks supply of either tobacco sticks or e-liquid |  |  |  |  |  | X |  | X | X | X |  | X | X | X | X | X | X |  |  |

**Table 2c:** Study Schedule of Procedures (Arm C – Never-Smokers)

| Procedure | Screening<br><up to 28<br>days before<br>V0> | Enrolment<br>(V0)<br>Day 1 | Baseline<br>(V1)<br>Day 14 ±3 | Week<br>12<br>(V2)<br>Day 90<br>±5 | Week<br>24<br>(V3)<br>Day<br>180 ±7 | Week<br>52<br>(V4)<br>Day<br>360 ±<br>10 | Week<br>104<br>(V5)<br>Day<br>540<br>±10 |
| --- | --- | --- | --- | --- | --- | --- | --- |
| Informed Consent | **X** |  |  |  |  |  |  |
| Socio-demographic data | **X** |  |  |  |  |  |  |
| Smoking Hx | **X** |  |  |  |  |  |  |
| Cigarette<br>Consumption<br>Check | **X** | **X** | **X** | **X** | **X** | **X** | **X** |
| Exhaled CO | **X** | **X** | **X** | **X** | **X** | **X** | **X** |
| Medical Hx | **X** |  |  |  |  |  |  |
| Assessment of<br>Pregnancy Status | **X** | **X** |  | **X** | **X** | **X** | **X** |
| Oral hygiene Hx | **X** | **X** | **X** | **X** | **X** | **X** | **X** |
| Periodontal<br>Examination | **X** |  |  |  |  |  |  |
| Eligibility Criteria<br>Check | **X** | **X** |  |  |  |  |  |
| Assignment to<br>Arm C |  | **X** |  |  |  |  |  |
| Tracker APP<br>installation |  | **X** |  |  |  |  |  |
| OHQOL<br>questionnaire |  | **X** | **X** | **X** | **X** | **X** | **X** |
| EQ VAS – QoL<br>questionnaire |  | **X** | **X** | **X** | **X** | **X** | **X** |
| Vital Signs (BP,<br>HR) |  | **X** | **X** | **X** | **X** | **X** | **X** |
| Weight, Height,<br>BMI |  | **X** |  | **X** | **X** | **X** | **X** |
| MGI |  | **X** | **X** | **X** | **X** | **X** | **X** |
| Plaque Score<br>Imaging |  | **X** | **X** | **X** | **X** | **X** | **X** |
| Tooth Stain<br>Assessment |  | **X** | **X** | **X** | **X** | **X** | **X** |
| Dental<br>Discoloration |  | **X** | **X** | **X** | **X** | **X** | **X** |
| Scaling &<br>Polishing |  | **X** |  |  |  |  | **X** |
| Safety reporting<br>(AE and SAE)/e-CRF |  | **X** | **X** | **X** | **X** | **X** | **X** |

Participants in Arm B will have the option to try and choose among a selection of either three e-liquids or three tobacco sticks (depending on the C-F NDS they have chosen). They will also be trained and instructed on how to correctly use their chosen C-F NDS.

Participants wishing to use a heated tobacco device (HTD) will receive one kit and supply of tobacco sticks of their choice enough to provide consumption in between supply visits; they will receive the number of tobacco sticks per day corresponding to the number of cigarettes smoked per day at baseline. Participants wishing to use a vaping product will receive one vaping kit and supply of e-liquids of their choice enough to provide consumption in between supply visits. Products will be supplied at each subsequent visit throughout the whole duration of the study according to Table 2b.

A prospective monitoring of cigarette consumption, C-F NDS use, and oral hygiene routine will be carried out throughout the study with the tracker APP. Moreover, participants in Arm B will be asked to return all empty, part-used, and unused consumables (tobacco sticks, e-cigarette cartridges, e-liquid refill bottles) at each study visit.

For all the further visits (Visit1-5) participants will have to agree to: refrain from scaling and polishing procedures; avoid modifying their own habitual oral hygiene (e.g .mouthwash, mouthrinse, interdental floss); refrain from flossing for at least 72 hours prior to each study visit; refrain from mouthrinsing for at least 24 hours prior to each study visit; refrain from toothbrushing for at least 2 hours prior to each study visit; refrain from eating and drinking (except water) for at least 2 hours before each visit; abstain from smoking for 2 hours before each visit; complete questionnaire via the automated Tracker App during the entire period of the study.

Baseline assessments will be performed at Visit 1, 14 days after Visit 0. Participants will be invited to attend four further visits (Visit 2-5) to undergo a series of assessment presented in the Schedule of Assessments (see Table 2a, Table 2b, Table 2c). Participant may be withdrawn from the study prematurely for the following reasons: 1) experiences an intolerable AE; 2) develops a concurrent disease which at the discretion of the Investigator no longer justifies the participant’s participation in this study; 3) decides to stop his/her participation; 4) becomes pregnant; 5) is uncooperative, including non-attendance. The overall duration of study participation for each participant will be a maximum of 568 +/-10 days.

### Objectives and Endpoints

The primary endpoint of the study is to evaluate changes in Modified Gingival Index (MGI) by Lobene ^(23)^ between baseline and end of study (i.e. 18 months timepoint) among participants who continue to smoke conventional cigarette and participants who switch from conventional cigarettes to C-F NDS.

Secondary endpoints include to assess within- and between-group (participants who continue to smoke conventional cigarettes, those who switch to C-F NDS and participants who never smoked) variations from baseline of the following study endpoints:

⍰ MGI (at 3, 6, 12, and 18 months)
⍰ Tooth Stains Assessment (at 3, 6, 12, and 18 months)
⍰ Dental Discolorations (at 3, 6, 12, and 18 months)
⍰ Plaque Score Imaging (at 3, 6, 12, and 18 months)
⍰ Oral Health Quality of Life (OHQOL) assessment (at 3, 6, 12, and 18 months)
• EuroQoL Visual Analog Scale (EQ VAS – QoL) assessment (at 3, 6, 12, and 18 months)

### Statistical Considerations

#### Sample Size Calculation

For a valid sample size calculation, a literature research was performed. Unfortunately, no data was available for such a study. However, data from studies with mouthrinse were available, see section References. Based on the studies from Cortelli, S. C., et al ^(24)^ and Lynch, M. C., et al ^(25)^ a mean MGI change of 0.08 with a standard deviation of 0.18 of the MGI seems relevant.

Nonetheless, in these studies higher mean MGI changes were observed, but in these studies an active treatment was given, which is not the case in this study. Thus, several scenarios with different mean MGI change and standard deviations will be applied. Moreover, it will be assumed that the data is normally distributed.

## Primary Endpoint

The following hypotheses will be tested with a one-sided independent t-Test: H0: µA ≤ µB versus H1: µA > µB

Where µA denotes the mean change MGI from baseline to after 18 months of regular smokers and the µB denotes the mean MGI from baseline to after 18 months of C-F NDS smokers.

### Sample Size Calculation

For the sample size calculation SAS software version 9.4 was used and several scenarios were applied. The results are presented in **Table A**. The following **Table A** presents the calculated sample sizes for the comparison of µA versus µB by a one-sided independent t-Test and a significance level of 5% and a power of 90% dependent on the expected MGI change and standard deviation.

**Table A:**
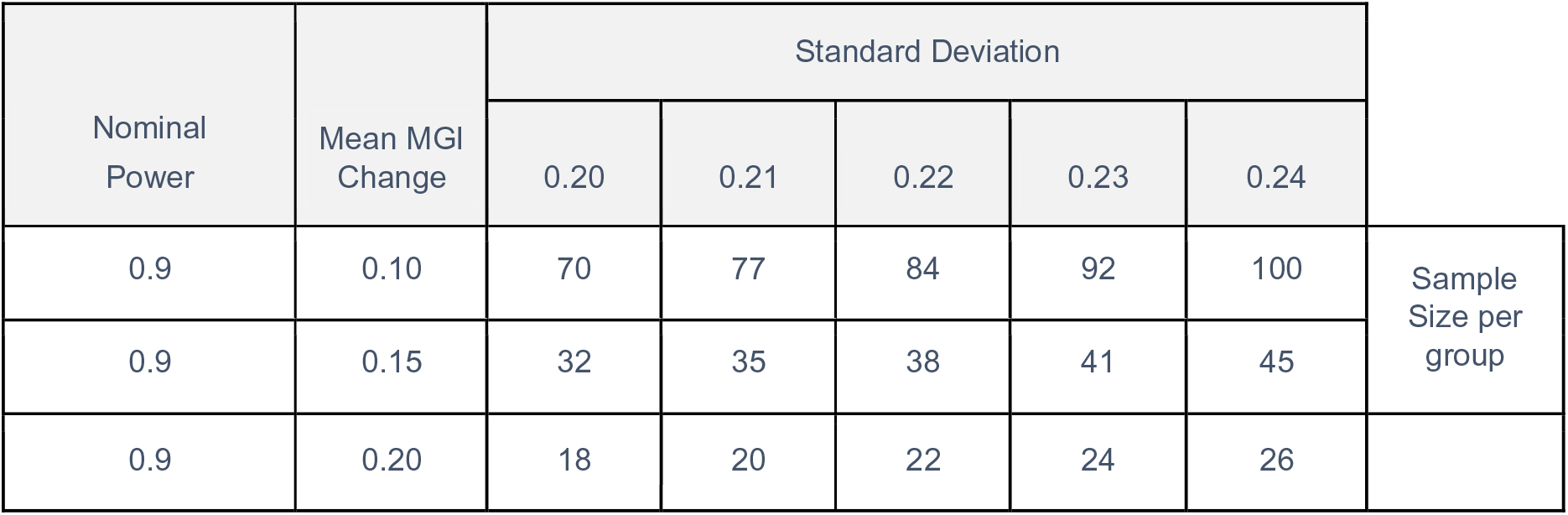
Summary table including the sample size per group depending on the mean MGI change and standard deviation

## Conclusion

Based on the studies from Cortelli, S. C., et al ^(24)^ and Lynch, M. C., et al ^(25)^ a mean MGI change of 0.08 with a standard deviation of 0.18 of the MGI seems relevant.

Nonetheless, in these studies higher mean MGI changes were observed, but in these studies an active treatment was given.

Since in the Smile Study no active treatment was given, a lower effect compared to the observed values needs to be assumed. Therefore, a mean MGI change of 0.1 with a standard deviation of 0.22 was chosen for the Smile study. Hence, a sample size of 84 per treatment group is required.

Nonetheless a dropout rate of 20% can be assumed for the two years observation phase ^(26, 27)^. Consequently, a sample size of 101 per treatment group is required.

In addition, it is estimated that about 25% of the switcher from conventional cigarette smokers to C-F NDS will abstain or substantially (≥ 90%) reduce smoking ^(28)^ and only these participants deliver valid results. Therefore, four times more participants should be recruited in the C-F NDS group (4*101 = 404).

These results in a total sample size of 101+404+101 = 606.

### Statistical Analyses

Statistical methods to be used in the analysis of this study will include: Descriptive statistic is defined as:

⍰ Descriptive presentation of N, mean, standard deviation, median, minimum, maximum and 95 % confidence limits will be given for (pseudo-) continuous data.
⍰ Descriptive presentation of N, mean, standard deviation, median, minimum and maximum will be given for categorical data. Further evaluations will be summarized in tables by counts and percentage of scores.

For primary endpoints:

⍰ Test of normality with Shapiro-Wilk on calculated values
  - In case of normal distribution: two-sided independent t-test
  - In case of no normal distribution: two-sided Mann-Whitney U test
⍰ Descriptive statistics
⍰ Linear Mixed Model (LMM) with the random factors center and Study Arms-by-center.

For secondary endpoints:

- For comparisons of each Study Arms for parameter MGI, Tooth Stains Assessment, Dental Discolorations, Plaque Score Imaging, OHQOL and EQ VAS - QoL:
  - Test of normality with Shapiro-Wilk on calculated values
    ⍰ In case of normal distribution: two-sided independent t-test
    ⍰ In case of no normal distribution: two-sided Mann-Whitney U test
- For comparisons of each assessment time for parameter MGI, Tooth Stains Assessment, Dental Discolorations, Plaque Score Imaging, OHQOL and EQ VAS - QoL:
  - Test of normality with Shapiro-Wilk on calculated values
    ⍰ In case of normal distribution: two-sided independent t-test
    ⍰ In case of no normal distribution: two-sided Mann-Whitney U test
- Accounting of Missing, Unused and Spurious Data

Only if one value of the parameter MGI is missing after Day 1, the last available observation will be carried forward (LOCF). The analysis will be performed on the substituted data. For secondary parameters no replacement of missing values will be applied.

## RESULTS

Patient recruitment will start in January 2021 and enrolment is expected to be completed by June 2021. Results will be reported between 2023 and in 2024.

## DISCUSSION

Although the negative effects of cigarette smoking on oral health and teeth appearance are well-known ^(29, 30)^, there is only limited data about the impact of combustion-free nicotine delivery systems (C-F NDS) such as e-cigarettes (ECs) and heated tobacco products (HTPs). In particular, there are no long-term studies assessing the impact on oral health and teeth appearance when substituting conventional cigarettes for these combustion-free alternatives. The SMILE Study was specifically designed to address these research questions. In particular, this study tests the hypothesis that avoiding exposure to cigarette smoke toxicants may translate into measurable amelioration in gingival response, dental plaque build-up, dental discoloration, and tooth staining in patients with gingivitis by comparing subjects with gingivitis who smoke tobacco cigarette with those who switch to using C-F NDS or subjects who never smoked.

Besides the obvious relevance to health and aesthetic concerns, the SMILE study seeks also to expand on regulatory science. Regulatory authorities (e.g. FDA in the US) recommend investigating oral health effects of novel tobacco/nicotine products to better understand their impact at individual and population level as well as exploring additional endpoints for the assessment of their short- and long-term effects in oral health studies ^(31)^. The SMILE Study will provide improved design knowledge for oral health switching studies and, most importantly, streamline new innovative cutting-edge technologies that can be used for future studies on existing and emerging tobacco/nicotine products.

The design of the study has many significant and groundbreaking characteristics. First, we can strengthen adherence to C-F NDS and optimize overall compliance with the study’s instructions by providing a wide variety of different products (reflecting the most popular of those commercially available). Participants can personalize a gratifying “nicotine experience” by selecting the C-F NDS of their preference/liking, thereby encouraging conversion to the latest technology and abstention from their own tobacco cigarettes ^(32, 33, 34, 35, 36)^. Please note that this element of choice has been missing in most studies. In addition, the results of the study will not be product-specific and unlikely to be limited in generalizability.

Second, the SMILE Study is being run in many different locations across the globe and so the challenge was to find a primary endpoint that could be measured by different operators in different sites, using a standardized measurement system that has been used previously and so has provenance. The index to be used had to be simple, reproducible and relatively easy to compare between different sites and different operators. Ideally the assessors should all be able to calibrate in the index to be used and the calibration results compared. The Modified Gingival Index (MGI) is a very widely used, industry standard index used to determine changes in the gingival health of volunteers in clinical studies ^(23, 37, 38)^. This index is simple non-invasive and reproducible and it is hoped to be possible to train different examiners in different locations to align in their assessments thus making the results comparable. Since the MGI is noninvasive there is no gingival probing, and it is thought to be easier to calibrate examiners and compare kappa scores. Although still subjective in nature the lack of probing removes one of the variables present in the using pressure to probe the gingivae. It is also though to afford greater sensitivity in determining therapeutic efficacy ^(39)^. The SMILE study will be the first one to investigate the long-term impact of smoking or CF-NDS intervention on MGI.

Third, the selection of secondary study endpoints is strategic, because it takes into account what drives smokers to turn to cleaner nicotine/tobacco products. This is particularly persuasive for young adults for whom a cardiovascular-cancer-respiratory risk-based narrative is either ineffective or even counterproductive and for whom concern about bad breath and teeth appearance (due to tooth discoloration and “tar” stains) may be a much more significant reason to refrain from smoking. Although several experimental papers comparing the effect of C-F NDS on tooth staining and discoloration have been published ^(40, 41, 42)^, the SMILE Study is the first to consider this powerful narrative of oral health in a clinical trial; we have therefore included innovative 21st century technologies for objective and consistent quantitation of dental discoloration (by calibrated spectrophotometry) and dental plaque changes (by digital imaging technology; QLF - Quantitative Light-induced Fluorescence) among the study endpoints to investigate if switching completely from cigarettes to C-F NDS can improve gum health, reduce bad breath and restore teeth appearance. Small-scale clinical trials have been recently conducted at the Center of Excellence for the Acceleration of Harm Reduction (CoEHAR), of the Catania University to confirm the validity and good reproducibility of QLF technology and calibrated spectrophotometry in current, former, and never smokers (unpublished data).

Fourth, maximizing the magnitude of observable changes of study endpoints is critically important. Given that several factors (including duration of smoking exposure, variations in oral hygiene practices, type of diet, level of alcohol consumption) can significantly affect assessments of gum health and dental esthetics, we reduced baseline variability by using data obtained after the visit during which scaling and polishing was carried out. By removing dental plaque, calculus and stains, we give all study participants the best possible oral health status (gum health and teeth appearance) at the beginning of the study. Considering that a 14-day interval is generally required to allow gingival restoration from tissue trauma caused by scaling and polishing ^(43)^, measurements that will be considered for this normalized baseline will be obtained 14 days after scaling and polishing. Any progressive change in gum health or teeth appearance will be compared to the reference data of this baseline. In addition to the inclusion of a full scale and polish at the beginning of the study in an attempt to maximize the magnitude of observable changes of study endpoints, it is also important to consider and that the length of the study is adequate and that the study population will have to be one which allows the possibility of measuring such a change. Although the long-term impact of smoking and smoking cessation on gingival health and teeth appearance has never been investigated in a prospective trial, the planned 18 months duration of SMILE study is deemed to be more than adequate for the detection of significant changes in study endpoints. Chronic periodontal disease is common in smokers and is an irreversible condition which may stabilize with active treatment ^(44)^, but is unlikely to actually improve with the simple measure of smoking cessation alone. Therefore, subjects with periodontitis will be excluded and only subjects with mild-moderate gingivitis will be recruited, as they are more likely to show measurable changes in study endpoints with continuing to smoke, smoking cessation and/ or the use of C-F NDS.

Fifth, personal oral hygiene and dietary patterns can significantly influence both primary and secondary study endpoints. Therefore, a standardized approach for oral hygiene is planned for the SMILE study; participants will be asked not to modify their routine oral hygiene regime for the whole duration of the study and to adhere to specific restriction criteria before each study visits to avoid confounding the data. In consideration of the variable impact of personal oral hygiene and dietary patterns in different countries, we included a third study arm of never-smokers as reference group. The challenge of measuring study endpoints by different operators in different sites, will be addressed by offering proper training and calibration.

Last but not least, compliance with the research protocol is important because it would decrease or nullify the anticipated improvements in study endpoints if cigarettes were not fully or largely replaced with C-F NDS. Participants will be reminded on the importance of adhering to their randomized product allocation and of abstaining from or substantially reducing the daily consumption of cigarettes (by at least 90% of their regular cigarette smoking at baseline) at every study visit. A significant feature of switching studies is close reporting of cigarette consumption and/or C-F NDS use; participants will record the consumption of cigarettes and use of C-F NDS on each visit in their study diary. In addition, subjects will be asked to return all empty, part-used, and unused consumables.

Throughout the study, smoking and C-F NDS usage will also be monitored via APP technology. The specially developed tracker APP for the SMILE study will also allow monitoring and verification of any non-compliance with the protocol as well as the oral health status of participants. Of note, non-compliance with study items is an interesting outcome in itself (particularly in consideration of the wide selection of different products offered in the study). Although compliance with this study is not expected to be significantly different compared to other comparable studies, our power calculations are overestimated to take account of a 75% non-compliance rate. The C-F NDS population would therefore be overrepresented by employing four times the number of subjects in the C-F NDS category (i.e., for every patient randomized in the control population, four will be randomized in the C-F NDS population).

## CONCLUSION

The SMILE study will be the first to measure oral health impact of smoking and smoking abstinence prospectively in the long term. The data generated will contribute to the existing knowledge base regarding the influence of smoking on oral health. In addition, the findings of the study may provide useful insights into the role of C-F NDS as a potentially cleaner nicotine alternative to tobacco smoking with the possibility of gains in oral health, especially for those smokers who are much more concerned about bad breath and the appearance of their teeth. Such a strong oral health narrative may have important implications for reducing the overall smoking burden.

The development of periodontal disease is correlated with several parameters measured in this study (including MGI and dental plaque build-up), and thus a more general impact on overall human health is also expected from the results of this study, given that progression of chronic gingivitis into periodontitis will subsequently increase the risk of cardiovascular disease ^(4, 5)^.

## Data Availability

Results will be reported between 2023 and in 2024

